# Sex Differences in the Association between Skeletal Muscle Energetics and Perceived Physical Fatigability: The Study of Muscle, Mobility and Aging (SOMMA)

**DOI:** 10.1101/2024.05.25.24307934

**Authors:** Emma L. Gay, Paul M. Coen, Stephanie Harrison, Reagan E. Garcia, Yujia (Susanna) Qiao, Bret H. Goodpaster, Daniel E. Forman, Frederico G. S. Toledo, Giovanna Distefano, Philip A. Kramer, Sofhia V. Ramos, Anthony J. A. Molina, Barbara J. Nicklas, Steven R. Cummings, Peggy M. Cawthon, Russell T. Hepple, Anne B. Newman, Nancy W. Glynn

## Abstract

Greater perceived physical fatigability and lower skeletal muscle energetics are predictors of mobility decline. Characterizing associations between muscle energetics and perceived fatigability may provide insight into potential targets to prevent mobility decline. We examined associations of *in vivo* (maximal ATP production, ATPmax) and *ex vivo* (maximal carbohydrate supported oxidative phosphorylation [max OXPHOS] and maximal fatty acid supported OXPHOS [max FAO OXPHOS]) measures of mitochondrial energetics with two measures of perceived physical fatigability, Pittsburgh Fatigability Scale (PFS, 0-50, higher=greater) and Rating of Perceived Exertion (RPE Fatigability, 6-20, higher=greater) after a slow treadmill walk. Participants from the Study of Muscle, Mobility and Aging (N=873) were 76.3±5.0 years old, 59.2% women, and 85.3% White. Higher muscle energetics (both *in vivo* and *ex vivo*) were associated with lower perceived physical fatigability, all p<0.03. When stratified by sex, higher ATPmax was associated with lower PFS Physical for men only; higher max OXPHOS and max FAO OXPHOS were associated with lower RPE fatigability for both sexes. Higher skeletal muscle energetics were associated with 40-55% lower odds of being in the most (PFS≥25, RPE Fatigability≥12) vs least (PFS 0-4, RPE Fatigability 6-7) severe fatigability strata, all p<0.03. Being a woman was associated with 2-3 times higher odds of being in the most severe fatigability strata when controlling for ATPmax but not the *in vivo* measures (p<0.05). Better mitochondrial energetics were linked to lower fatigability and less severe fatigability in older adults. Findings imply that improving skeletal muscle energetics may mitigate perceived physical fatigability and prolong healthy aging.

## Introduction

Perceived fatigability is a clinically useful indicator of an individual’s vulnerability to fatigue because it captures the effort individuals use to perform standard activities.^1,2^ Perceived physical fatigability is measured via questionnaire or immediately following a standardized physical task. Since perceived fatigability contextualizes whole body fatigue to the level of defined activity/task (i.e., intensity and duration) with which the fatigue is associated, it is a more sensitive measure of the extent to which an individual is limited by fatigue.^3–5^ Fatigability’s prevalence in older adults ranges from 20% to 89.5%, and is lower among men than women, with sex differences widening with age from 5% (age 60-69 years) to 21% (≥90 years).^6^ Greater perceived physical fatigability has been associated with deleterious health outcomes related to aging including: worse physical performance, mobility decline, impaired cognition, and all-cause mortality.^1,6–10^

Skeletal muscle mitochondria are essential energetic drivers of locomotion and mobility.^11–13^ Oxidative phosphorylation (OXPHOS) in the mitochondria of skeletal muscle is the primary source of adenosine triphosphate (ATP) for muscle contraction.^14^ Skeletal muscle mitochondrial energetics can be measured *in vivo* via maximal ATP production (ATPmax) and *ex vivo* via mitochondrial respiration of myofibers. New evidence from the Study of Muscle, Mobility and Aging (SOMMA) revealed sex differences in skeletal muscle energetics, where *in vivo* and *ex vivo* muscle energetics were significantly higher in men compared to women.^15^ Furthermore, substrate utilization during exercise may differ by sex with women reported to have a greater reliance on fatty acid oxidation to produce ATP, while men have a greater reliance on carbohydrate driven oxidation.^16,17^ Thus, *ex vivo* respiratory measures that capture ATP production from both fatty acid oxidation and carbohydrates may distinguish key physiological differences that determine sex-specific clinical domains.

Two prior studies have found associations between higher Rating of Perceived Exertion Fatigability (RPE Fatigability) and lowered capacity for OXPHOS, as measured via ATPmax.^18,19^ Thus, a potential driver of perceived fatigability may be inadequate energy supply for sustained muscle contraction and movement due to lower mitochondrial energy production. However, these studies lacked specific assessments of mitochondrial respiration and electron transport chain function. Additionally, Qiao et al., (2023) recently showed in the Study of Muscle, Mobility and Aging (SOMMA) that lower ATPmax, maximal complex I&II-supported OXPHOS (max OXPHOS), and maximal ETS capacity were associated with more severe performance fatigability (i.e., slowing down during a usual-paced 400m corridor walk).^20^ None of these three studies explored sex differences.

Mitochondrial energetics are known to decline with age, differ by sex and are strongly associated with slower gait speed and impaired physical function.^11,21,22^ Furthermore, greater perceived physical fatigability, a known prognostic indicator of mobility decline, is more prevalent in women than men.^5,6,9^ Thus, characterizing sex differences in the association between skeletal muscle energetics and perceived physical fatigability in a large cohort of older adults could provide novel insight into the potential role that mitochondrial energetics may play in aging and mobility decline (Figure 1). Therefore, the aim of our study was to extend prior work by examining sex differences in the association of mitochondrial energetics measured by spectroscopy and high resolution respirometry with perceived physical fatigability in a large well-characterized cohort of older adults. We hypothesized that associations of ATPmax and max OXPHOS with perceived physical fatigability would be stronger in men than women, while associations between maximal carbohydrate and fatty acid supported oxidation (max FAO OXPHOS) and perceived physical fatigability would be stronger in women than men. Additionally, we hypothesized that skeletal muscle energetics would have stronger associations with a perceived fatigability questionnaire that assessed multiple activities of various intensities and durations compared to perceived physical fatigability assessed after a slow walking task.

**Figure 1.**
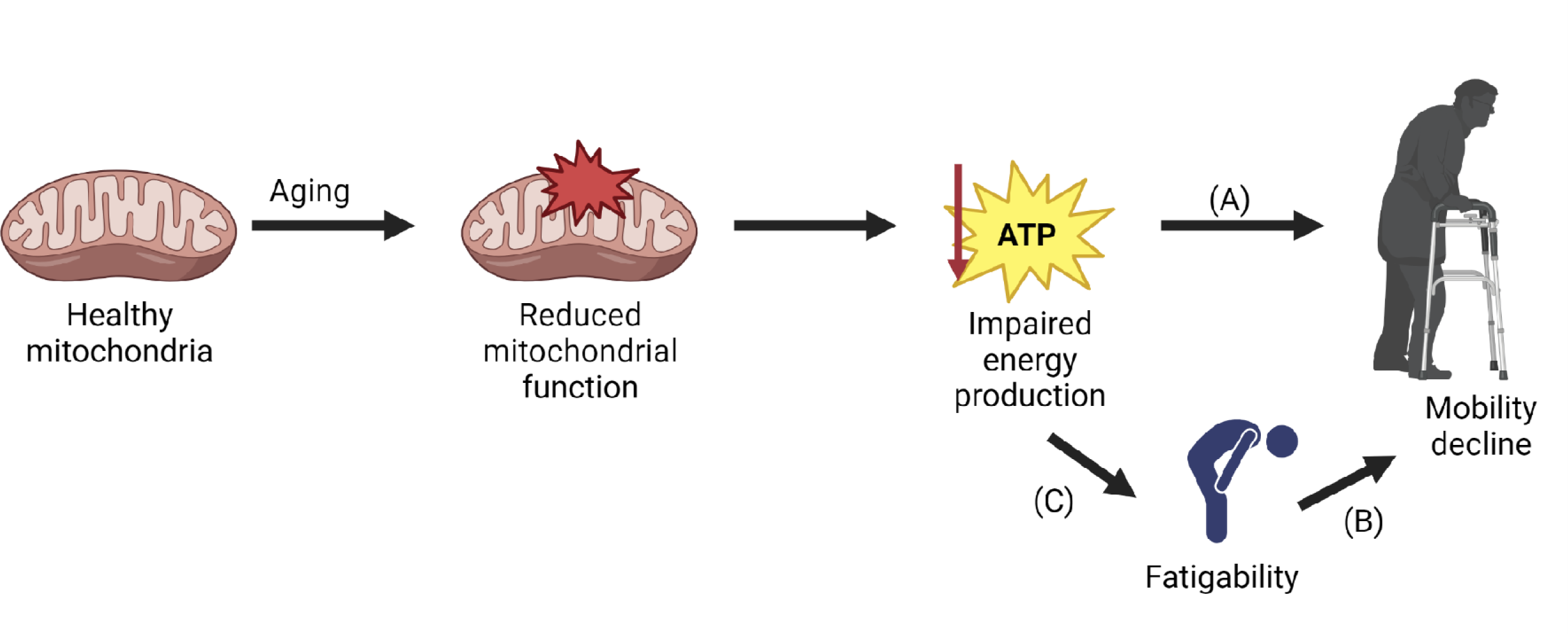
A Conceptual Model for the Association between Skeletal Muscle Energetics, Fatigability, and Mobility Decline As an individual ages, mitochondrial function declines, leading to impaired energy production. The association between skeletal muscle energetics and mobility decline is established (A). Fatigability is a known predictor of future mobility decline (B); however, the association between impaired energy production and fatigability in older adults is unknown (C). Understanding this association can inform potential future intervention strategies aimed at reducing fatigability to subsequently slowing mobility decline to improve healthy aging. Figure produced using BioRender.

## Methods

### Study Sample

A total of 879 community-dwelling adults aged ≥ 70 years were recruited for the Study of Muscle, Mobility and Aging (SOMMA, https://sommaonline.ucsf.edu) at the University of Pittsburgh and Wake Forest University School of Medicine between April 2019 and December 2021, as previously described.^23^ SOMMA provides the foundation for discoveries in the biology of human aging, mobility decline and other age-related phenotypes. Briefly, individuals were excluded at the time of telephone screening if they reported an inability to walk one-quarter of a mile or climb a flight of stairs; had a body mass index (BMI) ≤ 40 kg/m^2^; had an active malignancy or dementia; or any medical contraindication to biopsy or magnetic resonance imaging (MRI). Individuals were eligible to participate if they were willing and able to complete a skeletal muscle biopsy and undergo MRI and magnetic resonance spectroscopy (MRS). Lastly, participants must have been able to complete a usual-paced 400m walk within 15 minutes; those who appeared as they might not be able to complete the 400m walk at the in-person screening visit completed a 4m walk to ensure their walking speed was ≥ 0.6 m/s. All participants provided written informed consent, and SOMMA was approved by the Western IRB-Copernicus Groups Institutional Review Board (WCG-IRB; study number 20180764) as the single IRB for all participating sites.

### Skeletal Muscle Energetics

#### Maximal ATP Production (ATPmax)

Mitochondrial ATPmax was quantified using ^31^P MRS to measure the rate of phosphocreatine (PCr) regeneration following a short bout of exercise. This is a functional measure of in vivo ATP production via OXPHOS.^12^ SOMMA participants were instructed to lie supine with their lower leg strapped so the right knee had 20-30° of flexion. A 3 Telsa MRI scanner (Siemens Medical System – Prisma at Pittsburgh site or Skyra at Wake Forest site) using a 12” dual-tuned, surface radiofrequency (RF) coil (PulseTeq, Limited) placed over the right distal lateralis was used to collect ^31^P spectra. Participants performed two bouts of isometric knee extension against the resistance of an ankle strap. The first bout was as hard and fast as possible for 30 seconds, while the second was adjusted for duration of muscle contraction (18-36 seconds) based on the first bout the ensure 30-50% PCr depletion, which was required for a high signal to noise ratio, defining full PCr recovery without acidosis (pH < 6.8).^24^ PCr recovery rate after exercise was fit and the time-constant of the mono-exponential fit (tau) was used to calculate ATPmax.^25,26^ Previous work demonstrated good reproducibility of ATPmax (r=0.92) between same-day repeat scans.^18^ In SOMMA, the mean coefficient of variation for duplicate measures of ATPmax was 9.9% across clinic sites.^13^

#### Skeletal muscle biopsy collection and processing

The skeletal muscle biopsy was taken from the right vastus lateralis after a 12-hour fast and limited exercise for 48-hours prior to the procedure. Participants rested in a supine position for at least 15 minutes before the biopsy. The biopsy specimens were obtained under local anesthetic (1% or 2% lidocaine), about 15cm above the patella using a 5mm or 6mm Bergstrom-style core biopsy needly with suction. Muscle tissue was immediately blotted dry and trimmed of visible adipose and connective tissue. A portion (∼20mg) of the specimen was placed in a biopsy preserving solution for high-resolution respirometry, as previously described.^12^ Myofiber bundles (∼2-3mg) were prepared by gently separating the fibers using fine-point tweezers. The bundles were then chemically permeabilized in saponin for 30 minutes while rocking on ice, washed twice with mitochondrial respiration media (MiR05) for 10 minutes each, and then wet-weight was determined on an analytic balance (Mettler Toledo, Columbus, OH). Fiber bundle preparation was described in detail elsewhere.^13^

After weighing, the permeabilized myofiber bundles were placed into Oxygraph-2K respirometer chambers (O2K, Oroboros Instruments, Austria). Assays were run at 37°C within a specific range of O_2_ concentration (400-200µM). According to the assay protocol 1, the following substrates were added in sequential steps to assess maximal complex I&II supported OXPHOS at state 3 respiration (max OXPHOS): (1) pyruvate (5mM) and malate (2mM); (2) ADP (4.2mM), (3) Cytochrome c (10μM); (4) glutamate (10mM); and (5) succinate (10mM). For assay protocol 2, the following substrates were added in sequential steps to assess maximal carbohydrate and FAO supported OXPHOS (max FAO OXPHOS): (1) palmitoylcarnitine (25μM) and malate (2mM); (2) ADP (4mM); (3) glutamate (10mM) and Cytochrome c (10μM); and (4) succinate (10mM). Integrity of the outer mitochondrial membrane was tested with Cytochrome c, and any sample with an increase in respiration greater than 15% was excluded from analysis. Steady-state oxygen flux was normalized to the fiber bundle wet weight using DatLab 7.4 software. The mean coefficient of variation for duplicate measures of max OXPHOS was 11.5% across both sites.^13^

### Perceived Fatigability Measures

#### Pittsburgh Fatigability Scale (PFS)

The PFS, a validated 10-item measurement of perceived physical and mental fatigability,^3,27^ was self-administered prior to the baseline clinic visit. Participants rated the physical and mental fatigue that they expected or imagined they would experience immediately after completing activities of various intensity and duration on a scale from 0 (“no fatigue”) to 5 (“extreme fatigue”). Activities included in the PFS: leisurely walk for 30 minutes, brisk of fast walk for 1 hour, light household activity for 1 hour, heavy gardening or outdoor work for 1 hour, watching TV for 2 hours, sitting quietly for 1 hour, moderate-to-high-intensity strength training for 30 minutes, participating in a social activity for 1 hour, hosting a social event for 1 hour, and high-intensity activity for 30 minutes. PFS scores for physical and mental fatigability separately were summed across the 10-items (range 0 to 50, higher scores indicating greater fatigability). Scores were imputed for participants missing 1-3 items.^28^ For this analysis, only the PFS Physical subscale was included. An established cut-point of ≥15 classified participants as having more severe perceived physical fatigability.^5,9^ Further, PFS Physical severity strata were defined as 0-4, 5-9, 10-14, 15-19, 20-24, and ≥ 25.^5,9^

#### Rating of Perceived Exertion (RPE Fatigability)

During Day 2 of the baseline visit, participants completed a 3-phase treadmill cardiopulmonary exercise test (CPET) protocol.^29^ After Phase 1 (preferred walking speed test) and Phase 2 (maximal oxygen consumption test) participants had a 20-minute rest before proceeding to Phase 3, a slow walking speed treadmill test for 5 minutes at 0.67 m/s (1.5 mph) and 0% grade. Breath-by-breath VO_2_ and VCO_2_ were measured using a metabolic cart (Medgraphics Ultima Series, Medgraphics Corporation, St. Paul, MN) and facemask.^29^ Immediately after the end of the Phase 3 test, participants were asked to rate their perceived exertion using the Borg RPE scale (range 6-20, higher scores = greater RPE Fatigability).^30^ An establish cut-point of ≥10 classified participants with higher RPE fatigability.^5,31^ RPE Fatigability severity strata were defined as 6-7, 8-9, 10-11, and ≥ 12.^5,31^

### Other Variables of Interest

We obtained information on age, sex, and race (White, Black, Asian, Native American/Alaskan Native, Native Hawaiian/Pacific Islander, Multi-Racial, or Unknown) for each participant by self-reported questionnaire. Race was grouped for analysis as (1) White and (2) Black, Indigenous, and People of Color. We measured height (Harpenden stadiometers, Dyved, UK) and body mass (balance beam or digital scale) without shoes and with light clothing and calculated body mass index (BMI, weight [kg]/height [m^2^]). We combined information on self-reported history of physician diagnosed chronic health conditions and depressive symptoms to create the SOMMA multimorbidity index (0-11), a modification of the Rochester Epidemiology Project Multimorbidity Scale.^32^ Physical activity was measured by daily total activity count^33^ using the ActiGraph GT9X on the non-dominant wrist over a 7-day free-living period. The respiratory exchange ratio (RER) was calculated by dividing carbon dioxide output (VCO_2_) by the oxygen uptake (VO_2_) from the last recording at the end of the Phase 3 CPET test. An RER <0.7 indicates fatty acid oxidation, while an RER >1.0 indicates carbohydrate oxidation.^34,35^

### Statistical Analyses

Among enrolled participants (N=879), 873 participants completed the PFS Physical with scores imputed for 12 individuals. RPE Fatigability was available for 836 of the 873 participants.

Descriptive characteristics were reported as mean ± standard deviation (SD) or frequencies (n, %) for the overall sample and by sex. Differences by sex were examined using t-test for continuous and chi-square tests for categorical variables. The primary perceived physical fatigability study outcome was PFS Physical, whereas RPE Fatigability was used for replication. First, we examined Pearson correlations between muscle energetics and the perceived physical fatigability outcomes. Next, we generated linear regression models to evaluate the associations between skeletal muscle energetics and perceived physical fatigability (PFS Physical and RPE Fatigability in separate models) with progressive covariate adjustments to account for established and potential confounders based on prior literature.^7,13^ All muscle energetics variable units were standardized to 1 standard deviation intervals before analyses. Model 1 adjusted for technician/site, age, sex and race; Model 2 further adjusted for BMI; Model 3 further adjusted for total activity count; and the final model (i.e., fully adjusted) additionally adjusted for multimorbidity. Given our *a priori* hypothesis of sex differences in the association between skeletal muscle energetics and perceived physical fatigability, we stratified the fully adjusted model by sex, and formally tested for significant interactions (p ≤ 0.1). Standardized betas were generated for comparison between PFS Physical and RPE Fatigability models. Finally, multinomial logistic regression was used to examine the effect of 1 SD higher skeletal muscle energetics and sex on perceived physical fatigability severity, adjusting for age, sex, race, site/technician, BMI, total activity count, and multimorbidity. All analyses were performed in SAS 9.4.

## Results

### Participant Characteristics

Participants (N=873) were 76.3 ± 5.0 years old, 59.2% women, and 85.3% white (Table 1). Men were more likely to be white, less active and have a higher multimorbidity index than women, all p < 0.05. Age and body mass index (BMI) were similar between men and women (Table 1). Mean respiratory exchange ratio (RER) at the end of the 5 min treadmill walk was 0.80, indicative of a mixture of fatty acid and carbohydrate driven oxidation and did not significantly differ between men and women (Table 1). Mean PFS Physical score was nearly 3 points lower for men compared to women. The prevalence of higher perceived physical fatigability (PFS ≥ 15) was 54.1% overall (44.7% for men, 60.5% for women, p < 0.001). Mean ATPmax, mean max OXPHOS, and mean max FAO OXPHOS were higher for men compared to women, all p < 0.001 (Table 1).

**Table 1.**
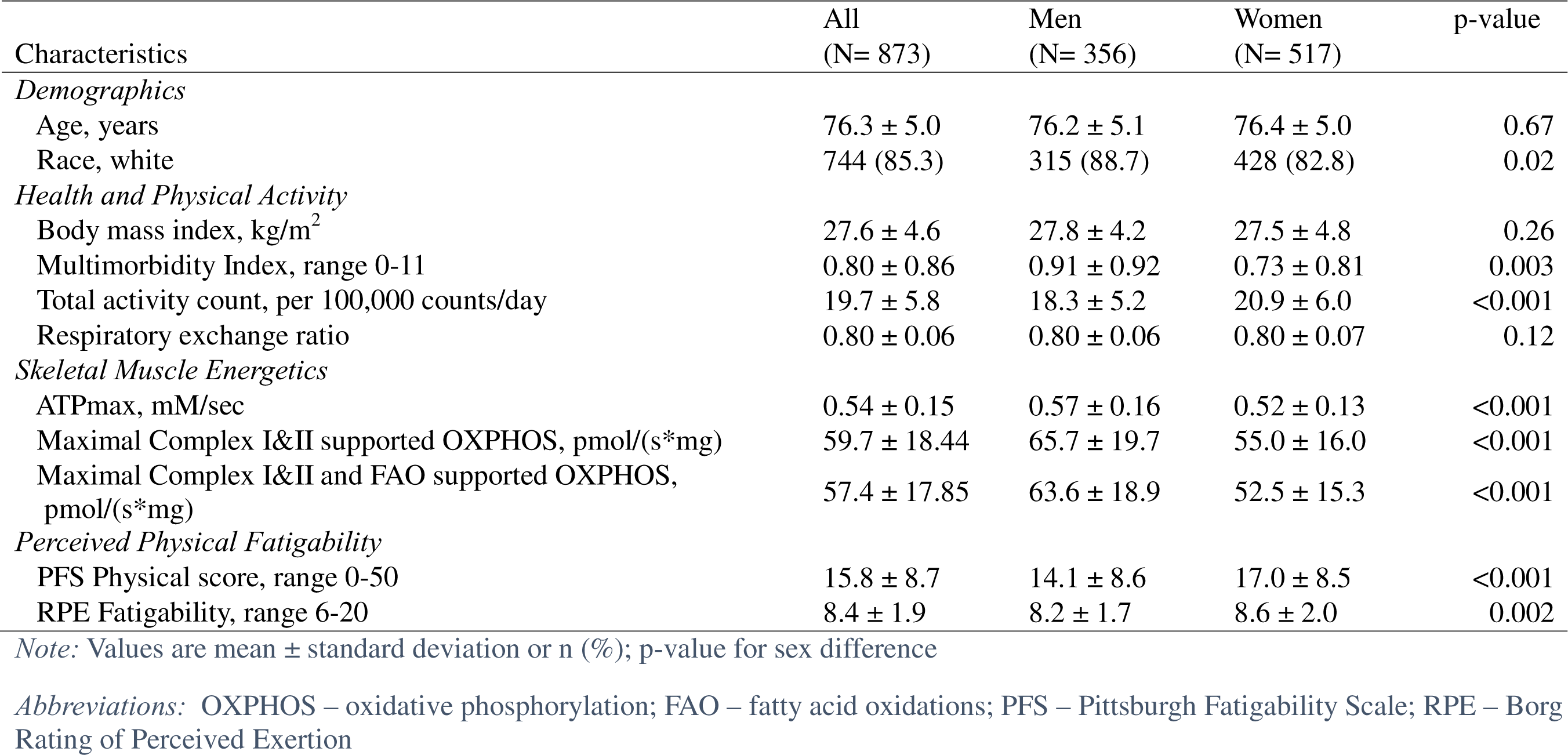
Sample Characteristics Overall and by Sex: The Study of Muscle, Mobility and Aging (SOMMA)

| Characteristics | All<br>(N= 873) | Men<br>(N= 356) | Women<br>(N= 517) | p-value |
| --- | --- | --- | --- | --- |
| *Demographics* |  |  |  |  |
| Age, years | 76.3 ± 5.0 | 76.2 ± 5.1 | 76.4 ± 5.0 | 0.67 |
| Race, white | 744 (85.3) | 315 (88.7) | 428 (82.8) | 0.02 |
| *Health and Physical Activity* |  |  |  |  |
| Body mass index, kg/m<sup>2</sup> | 27.6 ± 4.6 | 27.8 ± 4.2 | 27.5 ± 4.8 | 0.26 |
| Multimorbidity Index, range 0-11 | 0.80 ± 0.86 | 0.91 ± 0.92 | 0.73 ± 0.81 | 0.003 |
| Total activity count, per 100,000 counts/day | 19.7 ± 5.8 | 18.3 ± 5.2 | 20.9 ± 6.0 | <0.001 |
| Respiratory exchange ratio | 0.80 ± 0.06 | 0.80 ± 0.06 | 0.80 ± 0.07 | 0.12 |
| *Skeletal Muscle Energetics* |  |  |  |  |
| ATPmax, mM/sec | 0.54 ± 0.15 | 0.57 ± 0.16 | 0.52 ± 0.13 | <0.001 |
| Maximal Complex I&II supported OXPHOS, pmol/(s*mg) | 59.7 ± 18.44 | 65.7 ± 19.7 | 55.0 ± 16.0 | <0.001 |
| Maximal Complex I&II and FAO supported OXPHOS,<br>pmol/(s*mg) | 57.4 ± 17.85 | 63.6 ± 18.9 | 52.5 ± 15.3 | <0.001 |
| *Perceived Physical Fatigability* |  |  |  |  |
| PFS Physical score, range 0-50 | 15.8 ± 8.7 | 14.1 ± 8.6 | 17.0 ± 8.5 | <0.001 |
| RPE Fatigability, range 6-20 | 8.4 ± 1.9 | 8.2 ± 1.7 | 8.6 ± 2.0 | 0.002 |

*Note:* Values are mean ± standard deviation or n (%); p-value for sex difference
*Abbreviations:* OXPHOS – oxidative phosphorylation; FAO – fatty acid oxidations; PFS – Pittsburgh Fatigability Scale; RPE – Borg Rating of Perceived Exertion

### Pearson Correlations between Skeletal Muscle Energetics and Perceived Physical Fatigability

Lower ATPmax, max OXPHOS, and max FAO OXPHOS were all significantly, and moderately, correlated with higher PFS Physical score (Figure 2). Correlations between max OXPHOS and PFS Physical score were similar for men and women, however correlations for ATPmax and max FAO OXPHOS with PFS Physical score were stronger among men than women (Figure 3).

**Figure 2.**
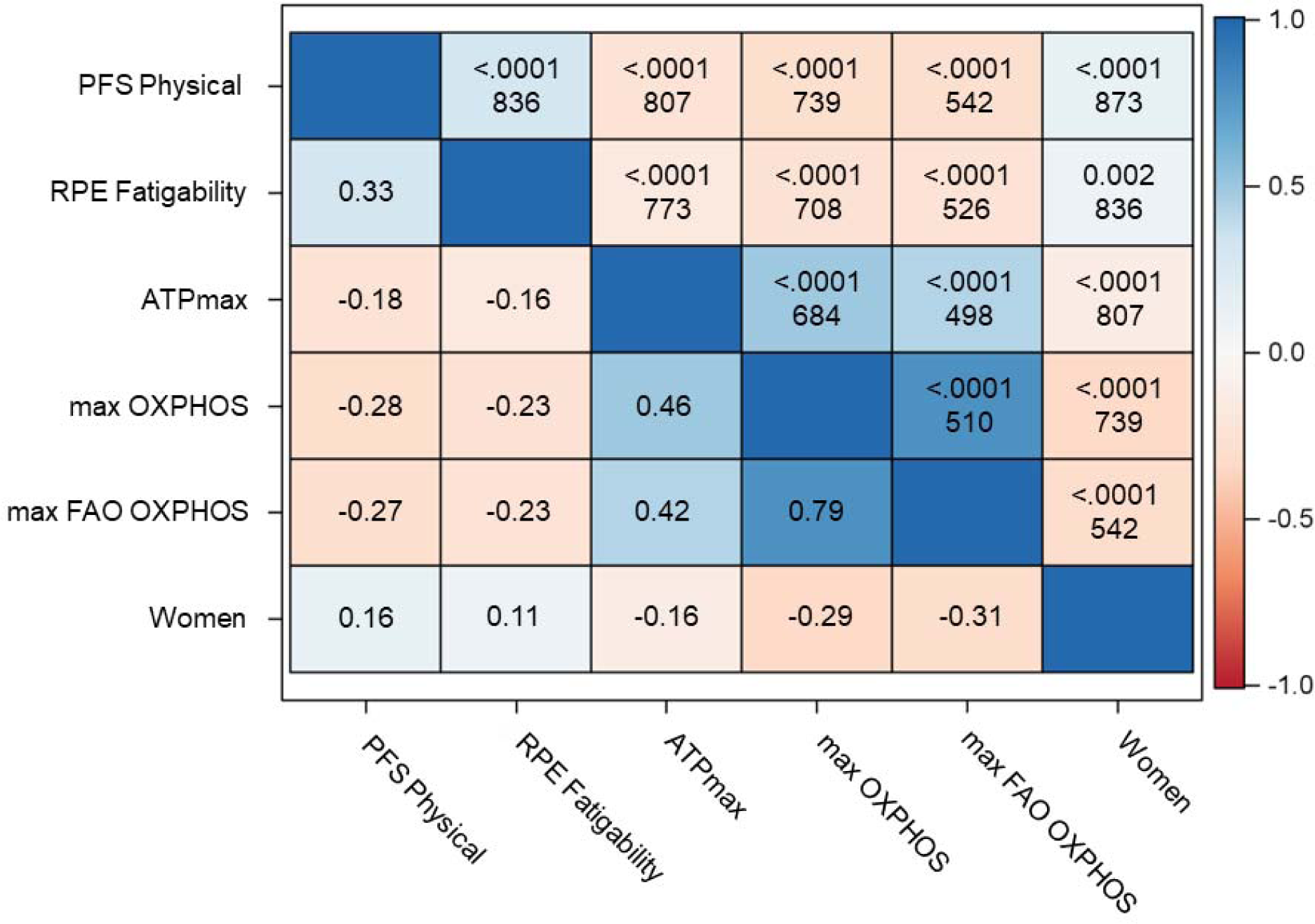
Pearson Correlation Matrix Between Skeletal Muscle Energetics and Perceived Physical Fatigability: The Study of Muscle, Mobility and Aging (SOMMA) *Abbreviations:* ATPmax (mM/sec); Max OXPHOS – Maximal Complex I&II Supported OXPHOS (pmol/(s*mg)); Max FAO OXPHOS – Maximal Complex I&II Supported FAO OXPHOS (pmol/(s*mg)); OXPHOS – oxidative phosphorylation; FAO – fatty acid oxidation; PFS – Pittsburgh Fatigability Scale; RPE – Borg Rating of Perceived Exertion

**Figure 3.**
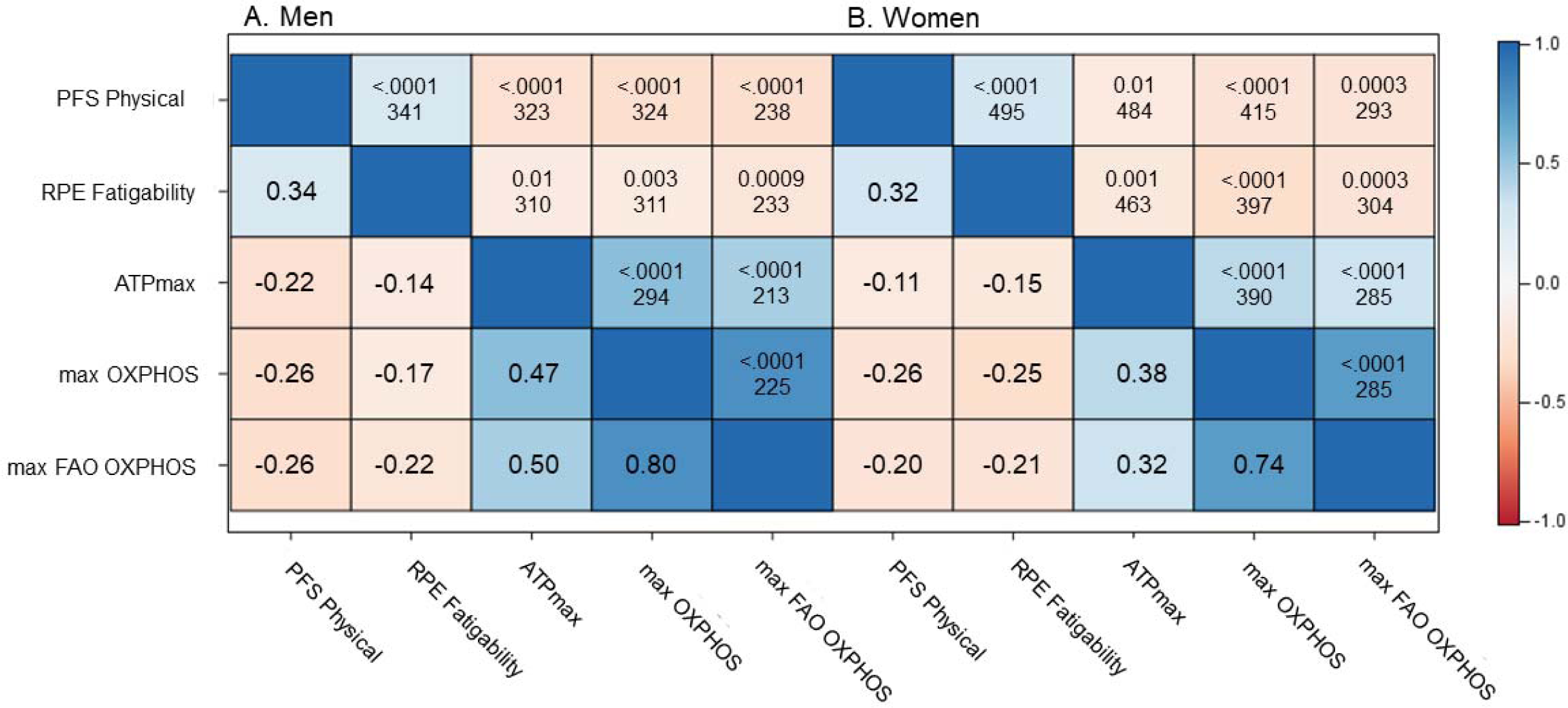
Pearson Correlation Matrices between Skeletal Muscle Energetics and Perceived Physical Fatigability Stratified by Sex: The Study of Muscle, Mobility and Aging (SOMMA) *Abbreviations:* ATPmax (mM/sec); Max OXPHOS – Maximal Complex I&II Supported OXPHOS (pmol/(s*mg)); Max FAO OXPHOS – Maximal Complex I&II Supported FAO OXPHOS (pmol/(s*mg)); OXPHOS – oxidative phosphorylation; FAO – fatty acid oxidation; PFS – Pittsburgh Fatigability Scale; RPE – Borg Rating of Perceived Exertion

### Associations between Skeletal Muscle Energetics and Perceived Physical Fatigability

The minimally adjusted model (Model 1) showed that higher skeletal muscle energetics were significantly associated with lower perceived physical fatigability on both measures (Supplemental Table 1). The addition of BMI (Model 2) slightly attenuated by 1-2% all associations between skeletal muscle energetics and perceived physical fatigability. Adding total activity count (Model 3) further attenuated by 0-4% all associations between skeletal muscle energetics and perceived physical fatigability (Supplemental Table 1).

For the fully adjusted ATPmax model that included age, sex, race, site, BMI, total activity count and multimorbidity index, each 1 standard deviation (SD) unit higher ATPmax was associated with approximately 1.0 point lower PFS Physical score, p = 0.003 (Table 2). When stratified by sex, each 1 SD unit higher ATPmax was associated with a 1.5 point lower PFS Physical score for men, p = 0.003, and a 0.4 point lower PFS Physical score for women, p = 0.3, p-interaction = 0.03 (Table 2).

**Table 2.** Linear Regression Models for Association of Skeletal Muscle Energetics on Perceived Physical Fatigability Overall and by Sex: The Study of Muscle, Mobility and Aging (SOMMA)

|  | PFS Physical score |  |  |  |  | RPE Fatigability |  |  |  |
| --- | --- | --- | --- | --- | --- | --- | --- | --- | --- |
| | unit | $\beta$ | $\beta$ | 95% CI | p-value | $\beta$ | $\beta$ | 95% CI | p-value |
| *Overall* |  |  |  |  |  |  |  |  |  |
| ATPmax | 0.15 | -0.11 | -0.95 | (-1.56, -0.34) | 0.003 | -0.08 | -0.16 | (-0.29, -0.03) | 0.02 |
| max OXPHOS | 18.44 | -0.20 | -1.71 | (-2.36, -1.05) | <0.001 | -0.13 | -0.25 | (-0.39, -0.10) | <0.001 |
| max FAO OXPHOS | 17.85 | -0.19 | -1.64 | (-2.41, -0.88) | <0.001 | -0.18 | -0.34 | (-0.51, -0.16) | <0.001 |
| *Men* |  |  |  |  |  |  |  |  |  |
|  |  | - |  |  |  |  |  |  |  |
|  |  | 0.17 |  |  |  |  |  |  |  |
| ATPmax | 0.16 | □ | -1.51 | (-2.49, -0.52) | 0.003 | -0.09 | -0.15 | (-0.36, 0.05) | 0.13 |
| max OXPHOS | 19.66 | -0.21 | -1.80 | (-2.75, -0.85) | <0.001 | -0.12□ | -0.20 | (-0.39, 0.00) | 0.05 |
| max FAO OXPHOS | 18.92 | -0.22 | -1.87 | (-2.98, -0.76) | 0.001 | -0.18□ | -0.31 | (-0.54, -0.07) | 0.01 |
| *Women* |  |  |  |  |  |  |  |  |  |
|  |  | - |  |  |  |  |  |  |  |
|  |  | 0.05 |  |  |  |  |  |  |  |
| ATPmax | 0.13 | □ | -0.41 | (-1.17, 0.36) | 0.30 | -0.08 | -0.16 | (-0.33, -0.01) | 0.07 |
| max OXPHOS | 15.97 | -0.19 | -1.58 | (-2.43, -0.73) | <0.001 | -0.16□ | -0.32 | (-0.51, -0.13) | 0.001 |
| max FAO OXPHOS | 15.31 | -0.17 | -1.43 | (-2.41, -0.45) | 0.004 | -0.20□ | -0.40 | (-0.63, -0.16) | 0.001 |

All models adjusted for age, sex (overall model only), race, site/technician, body mass index, total activity count, and multimorbidity index.
□ (0.03), □ (0.1), □ (0.1) denote significant interaction term for sex differences in skeletal muscle energetics
*Abbreviations:* ATPmax (mM/sec); Max OXPHOS – Maximal Complex I&II Supported OXPHOS (pmol/(s\*mg)); Max FAO OXPHOS – Maximal Complex I&II Supported FAO OXPHOS (pmol/(s\*mg)); OXPHOS – oxidative phosphorylation; FAO – fatty acid oxidation; PFS – Pittsburgh Fatigability Scale; RPE – Borg Rating of Perceived Exertion; $\beta$ – standardized beta coefficient; $\beta$ – beta coefficient; CI – confidence interval

For the fully adjusted max OXPHOS model, each 1 SD unit higher max OXPHOS was associated with a 1.7 point lower PFS Physical score, p < 0.001 (Table 2). When stratified by sex, each 1 SD unit higher max OXPHOS was associated with a 1.8 point lower PFS Physical score for men, p < 0.001, and a 1.6 point lower PFS Physical score for women, p < 0.001, p-interaction = 0.9 (Table 2).

Lastly for the fully adjusted model for max FAO OXPHOS, each 1 SD unit higher max FAO OXPHOS was associated with a 1.6 point lower PFS Physical score, p < 0.001 (Table 2). When stratified by sex, each 1 SD unit higher max FAO OXPHOS was associated with a 1.9 point lower PFS Physical score for men, p = 0.001, and a 1.4 point lower PFS Physical score for women, p = 0.004, p-interaction = 0.9 (Table 2).

### Associations of Skeletal Muscle Energetics with Perceived Fatigability Severity

Each 1 SD higher ATPmax was associated with a 40% lower odds of being in the most severe (≥ 25) PFS Physical severity strata compared to the least severe (0-4) after adjusting for age, sex, race, site/technician, BMI, total activity count, and multimorbidity (OR=0.60, 95% CI 0.44-0.82, Figure 4). Similar lower odds of being in the most severe fatigability severity strata were observed for max OXPHOS and max FAO OXPHOS (Figure 4). Women had 3-fold higher odds of being in the most severe (≥ 25) PFS Physical severity strata than the least severe (0-4) compared to men for ATPmax only after covariate adjustment (OR = 3.18, 95% CI 1.64-6.19, Figure 4). After adjusting for max OXPHOS and max FAO OXPHOS in separate models, women did not have a significantly higher odds of being in the most severe PFS Physical severity strata compared to men (Figure 4).

**Figure 4.**
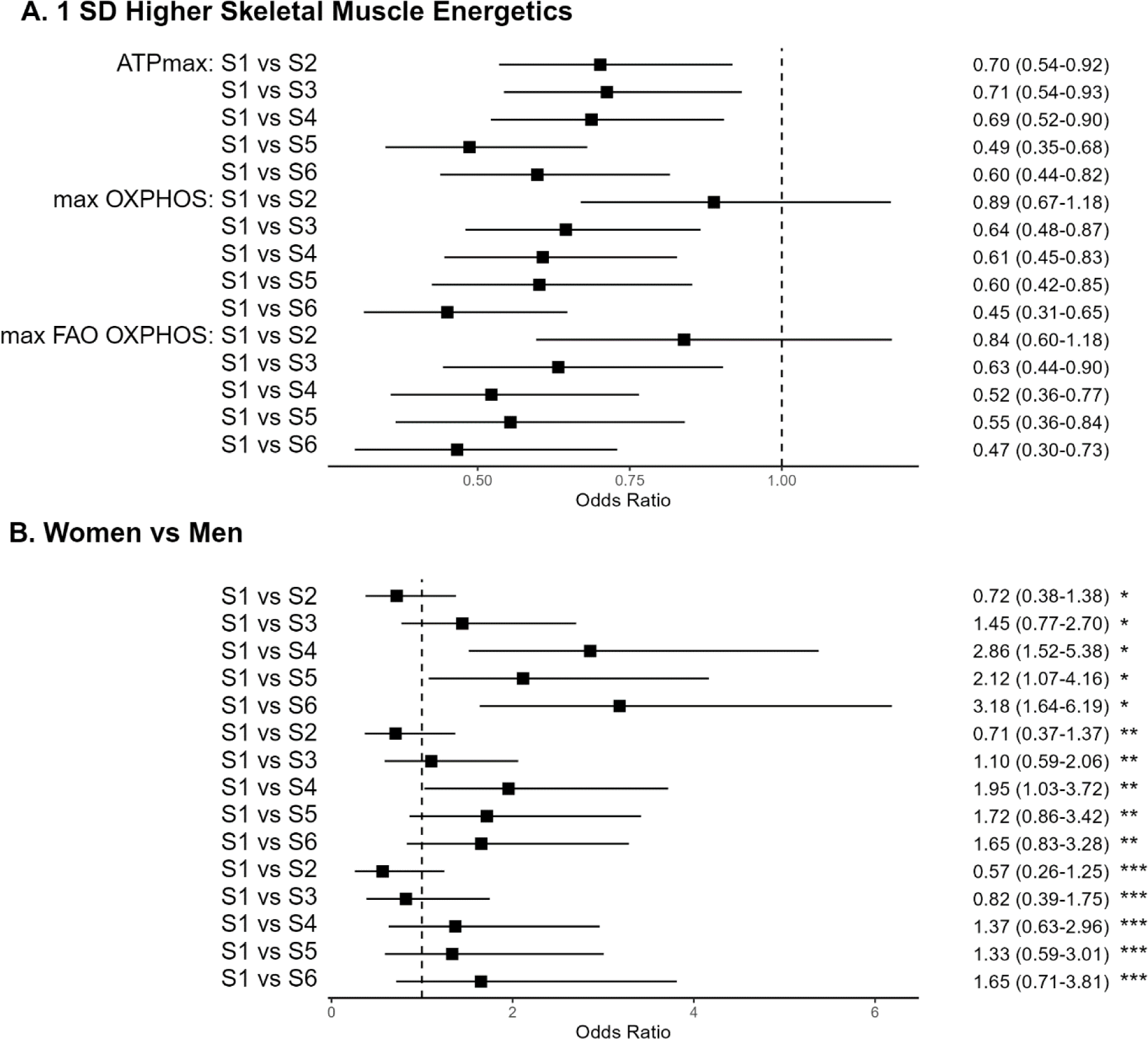
Multinomial Logistic Regression for the Association of Skeletal Muscle Energetics with PFS Physical Severity Strata Overall and Between Women and Men: The Study of Muscle, Mobility and Aging **PFS Severity strata:** S1 (0-4), S2 (5-9), S3 (10-14), S4 (15-19), S5 (20-24), S6 (≥ 25). Panel A. Odds of having more severe fatigability (i.e., S2-S6) with 1 standard deviation (SD) unit higher skeletal muscle energetics compared to the least severe (S1) adjusting for age, sex, race, site/technician, body mass index (BMI), total activity count, and multimorbidity. Panel B. Odds of being in a higher severity strata (i.e., S2-S6) for women compared to men adjusting for age, race, site/technician, BMI, total activity count, multimorbidity, and skeletal muscle energetics (*ATPmax, **max OXPHOS, ***max FAO OXPHOS). *Abbreviations:* ATPmax (mM/sec); Max OXPHOS – Maximal Complex I&II Supported OXPHOS (pmol/(s*mg)); Max FAO OXPHOS – Maximal Complex I&II Supported FAO OXPHOS (pmol/(s*mg)); OXPHOS – oxidative phosphorylation; FAO – fatty acid oxidation; PFS – Pittsburgh Fatigability Scale

### Replication with RPE Fatigability

The prevalence of higher RPE fatigability (RPE ≥ 10) was 20.2% overall (16.1% for men; 20.3% for women, p = 0.02), three-fold lower perceived fatigability than when measured using the PFS. In contrast to PFS Physical, correlations between max OXPHOS and FAO OXPHOS with RPE Fatigability were weaker among men than women, while for ATPmax, correlations with RPE Fatigability were similar between men and women (Figure 3). Overall, for the fully adjusted model, higher ATPmax, max OXPHOS, and max FAO OXPHOS were associated with lower RPE Fatigability, all p < 0.05 (Table 2). Counter to PFS Physical, there were no sex differences with ATPmax and RPE Fatigability, p-interaction = 0.8. However, unlike PFS Physical, for RPE Fatigability the sex × max OXPHOS and sex × max FAO OXPHOS interaction terms were significant, both p-interaction = 0.1 (Table 2). In the multinomial logistic regression model, similar significant associations were observed for odds of being in the most severe (≥ 12) vs least severe (6-7) RPE Fatigability severity strata as for PFS Physical severity strata (Figure 5). Additionally, similar associations were observed for women after adjusting for ATPmax, but not max OXPHOS and max FAO OXPHOS (Figure 5).

**Figure 5.**
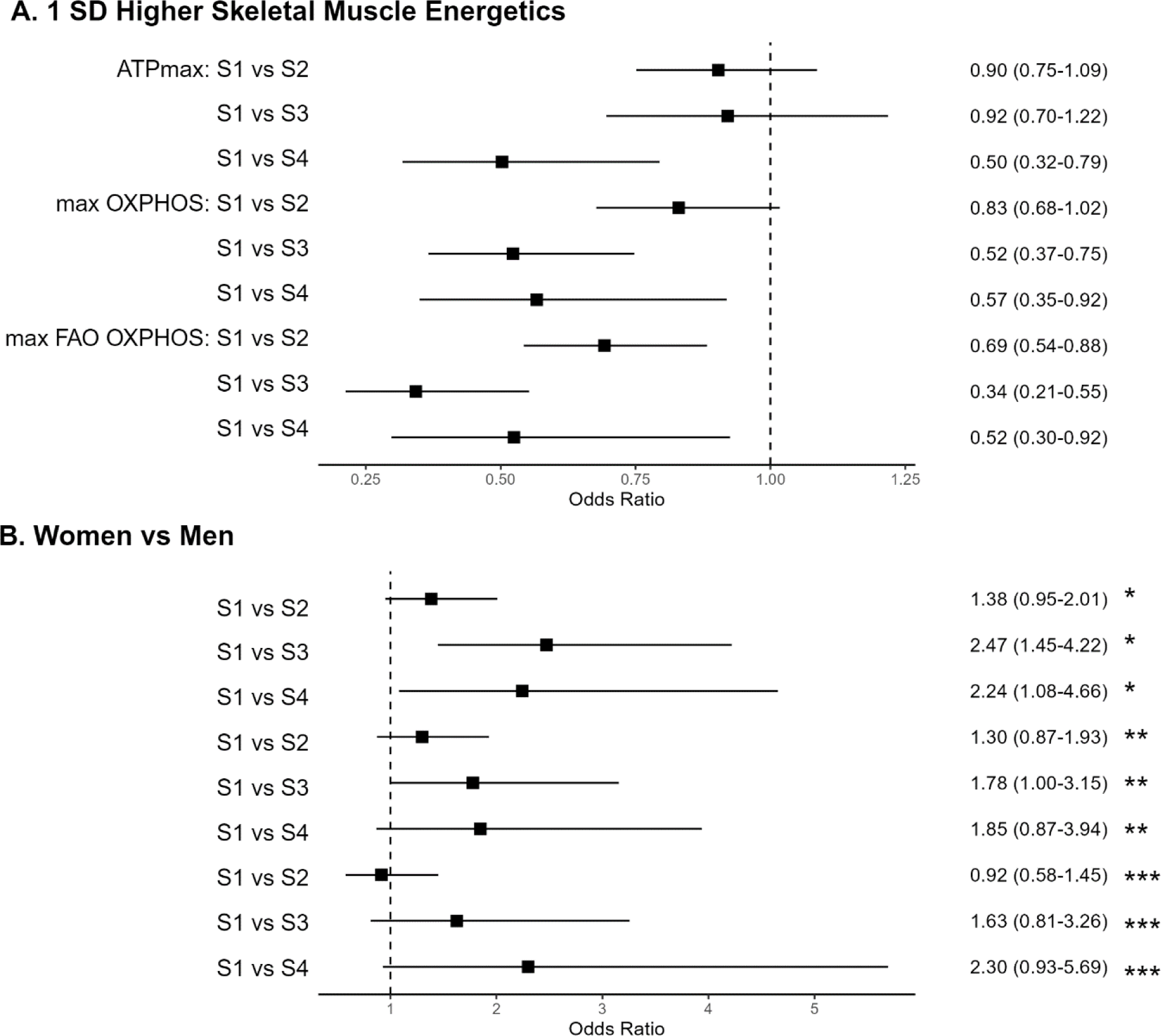
Multinomial Logistic Regression for the Association of Skeletal Muscle Energetics with RPE Fatigability Severity Strata Overall and Between Women and Men: The Study of Muscle, Mobility and Aging **RPE Fatigability strata:** S1 (6-7), S2 (8-9), S3 (10-11), S4 (≥ 12). Panel A. Odds of having more severe fatigability (i.e., S2-S4) with 1 standard deviation (SD) interval higher skeletal muscle energetics compared to the least severe (S1) adjusting for age, sex, race, site/technician, body mass index (BMI), total activity count, and multimorbidity. Panel B. Odds of being in a higher severity strata (i.e., S2-S4) for women compared to men adjusting for age, race, site/technician, BMI, total activity count, multimorbidity, and skeletal muscle energetics (*ATPmax, **max OXPHOS, ***max FAO OXPHOS). *Abbreviations:* ATPmax (mM/sec); Max OXPHOS – Maximal Complex I&II Supported OXPHOS (pmol/(s*mg)); Max FAO OXPHOS – Maximal Complex I&II Supported FAO OXPHOS (pmol/(s*mg)); OXPHOS – oxidative phosphorylation; FAO – fatty acid oxidation; RPE – Borg Rating of Perceived Exertion

## Discussion

To our knowledge, this is the first study to evaluate the association between skeletal muscle energetics, both *in vivo* and *ex vivo,* with perceived physical fatigability measured using both questionnaire and task-based methodologies. We found that higher skeletal muscle energetics were significantly associated with lower perceived physical fatigability in a large cohort of well phenotyped older adults, with some notable sex differences. Our findings support an important role for energetics in perceived physical fatigability, a common trait in older adults, and suggests skeletal muscle energetics is a likely therapeutic target for intervention to mitigate greater perceived physical fatigability.

For PFS Physical, the magnitude of association with ATPmax was 4 times greater in men than women, while max OXPHOS and max FAO OXPHOS associations were of a similar magnitude between the sexes. After adjustment for BMI, which affects muscle energetics and differs by sex,^36^ the association between ATPmax and PFS Physical was only significant in men and not women. The association of max OXPHOS and max FAO OXPHOS with RPE Fatigability was significantly modified by sex and stronger in women compared to men. This may be due to the low intensity of the slow walking task. Although women tend to prefer FAO substrates whereas men prefer carbohydrate substrates,^37^ the respiratory exchange ratio at the end of the 5 minute slow treadmill walk was not significantly different by sex (Table 1). Further, a greater capacity for max OXPHOS and max FAO OXPHOS was associated with lower RPE Fatigability in both sexes, but the association was stronger among women. This may reflect the fact that women are known to have a greater reliance on fatty acid oxidation to produce ATP during activity^37,38^ and found the slow walking task to be more fatiguing than men.

The prevalence of greater PFS Physical fatigability observed in the SOMMA cohort was approximately 5-10% higher compared to the Baltimore Longitudinal Study of Aging (BLSA) and the Long Life Family Study.^6,7,9^ The prevalence of higher RPE Fatigability in this cohort was comparable to the reported prevalence in BLSA.^6,30^ While both measures assess perceived physical fatigability anchored to specific tasks, the PFS was designed to evaluate fatigability for multiple activities across varying intensities (light to heavy) and duration, whereas RPE Fatigability was assessed directly after a 5-minute slow (0.67 m/s) treadmill walking task. Thus, the stronger associations of ATPmax and max OXPHOS with PFS Physical may reflect the changes in energy capacity needed to participate in real-world activities with higher energetic demands. Our results may reflect that older adults with higher mitochondrial oxidative capacity are better able to maintain their lifestyle with less symptoms of fatigue.

Our findings from multinomial logistic regression indicate that higher skeletal muscle energetics was associated with lower odds of being in a higher perceived physical fatigability strata. Interestingly, when controlling for ATPmax, women had over 2 times higher odds of being in the most severe physical fatigability strata compared to men. However, after controlling for max OXPHOS and max FAO OXPHOS in separate models, no significant differences between men and women were observed. Thus, for women OXPHOS and FAO pathways may be targets for intervention in order to reduce the higher prevalence of perceived physical fatigability experienced by women in order to stem its deleterious consequences, such as mobility decline, frailty, and mortality.^6^

Our results confirm and extend previous work associating ATPmax with perceived physical fatigability, as measured by RPE Fatigability. Our associations were of a similar magnitude to the prior work by both Santanasto et al. and Liu et al.^18,19^ Our novel work complements this existing knowledge by associating two *ex vivo* measures of skeletal muscle energetics, max OXPHOS and max FAO OXPHOS, with a globally used questionnaire-based measure of perceived physical fatigability, the PFS, which does not require special equipment or research staff to administer. We also address differences in these associations based upon sex.

There are some limitations as well as strengths to our work. First, 85% of the sample was Caucasian, dampening the generalizability of our results. There is evidence that mitochondrial energetics may differ by race/ethnicity.^39^ Second, the respiratory measures of OXPHOS do not account for potential changes in efficiency in the amount of respiration to needed support a given level of phosphorylation of ADP to form ATP, noting that a reduced mitochondrial phosphorylation efficiency with aging has been observed in older adults.^40^ Third, this study assumes a causality direction from skeletal muscle energetics to perceived fatigability and mobility. However, it is possible that reduced mobility leads to deconditioning which in turn causes higher fatigability and lower muscle energetics. Finally, respirometry measures were unable to determine if observed associations were due to electron transport chain activity or changes in mitochondrial content. Although the SOMMA cohort tended to be healthier than the general population, we enrolled individuals across a wide range of physical function, included comprehensive *in vivo* and *ex vivo* assessments of skeletal muscle energetics, and assessed outcomes with two widely used validated measures of perceived physical fatigability, all bolstering the generalizability of our results.

In conclusion, higher skeletal muscle energetics were significantly associated with lower perceived physical fatigability in older adults, and the associations were stronger for ATPmax and max OXPHOS with PFS Physical and for max FAO OXPHOS with RPE Fatigability. The association between ATPmax and PFS Physical was modified by sex, with significant associations observed for men, but not women, although the reasons for this are not clear. The associations of max OXPHOS and max FAO OXPHOS with RPE Fatigability were also modified by sex, but in this case stronger associations were observed for women versus men. Evaluation of mitochondrial content is needed to understand the degree to which these associations are influenced by intrinsic differences in electron transport chain activity versus changes in mitochondrial content. Previous research has demonstrated that as individuals age, men are more likely to have an intrinsic respiratory defect,^41^ while women are more likely to have a reduction in mitochondrial content.^42^ Longitudinal data are needed to further understand the temporal association between skeletal muscle energetics and perceived physical fatigability and the impact of sex. Increasing skeletal muscle energetics via physical activity or novel therapeutics may be a potential target for future interventions aimed at reducing fatigability to subsequently slowing mobility decline to improve healthy aging and may provide opportunities to tailor care to address sex-specific clinical differences.

## Data Availability

All data produced in the present study are available upon reasonable request to the author. SOMMA data is publicly available by request at https://sommaonline.ucsf.edu/.

## Acknowledgements

The Study of Muscle, Mobility and Aging is supported by funding from the National Institute on Aging (R01 AG 059416). Study infrastructure support was funded in part by NIA Claude D. Pepper Older American Independence Centers at University of Pittsburgh (P30 AG024827) and Wake Forest University (P30 AG021332) and the Clinical and Translational Science Institutes, funded by the National Center for Advancing Translational Science, at Wake Forest University (UL1 TR001420). Additionally, the NIA Claude D. Pepper Older Americans Independence Center, Research Registry and Developmental Pilot Grant (P30 AG024827), and the Intramural Research Program, National Institute on Aging supported N.W.G to develop the Pittsburgh Fatigability Scale.

## Data and Code Availability

SOMMA data is publicly available by request at https://sommaonline.ucsf.edu/. The SAS code used to analyze the data is available upon request.

## Authors’ contributions

Ms. Gay, Ms. Harrison and Dr. Glynn had full access to all of the data for the manuscript and take responsibility for the integrity of the data and accuracy of the data analyses. Ms. Gay wrote the first draft under Dr. Glynn’s supervision. All authors: interpretation of data, critical revision of manuscript for important intellectual content. All authors read and approved the submitted manuscript.

---

**Supplemental Table 1.** Linear Regression Models for the Association between Skeletal Muscle Energetics and Perceived Physical Fatigability: The Study of Muscle, Mobility and Aging (SOMMA)

|  | Model 1 |  |  |  |  | Model 2 |  |  |  | Model 3 |  |  |  |
| --- | --- | --- | --- | --- | --- | --- | --- | --- | --- | --- | --- | --- | --- |
| | unit | $\beta$ | $\beta$ | 95%CI | p-value | $\beta$ | $\beta$ | 95%CI | p-value | $\beta$ | $\beta$ | 95%CI | p-value |
| ***PFS Physical*** |  |  |  |  |  |  |  |  |  |  |  |  |  |
| ATPmax | 0.15 | -0.16 | -1.39 | (-1.99, -0.79) | <0.001 | -0.15 | -1.27 | (-1.86, -0.68) | <0.001 | -0.11 | -0.93 | (-1.55, -0.32) | 0.003 |
| max OXPHOS | 18.44 | -0.23 | -2.02 | (-2.67, -1.37) | <0.001 | -0.21 | -1.85 | (-2.49, -1.20) | <0.001 | -0.21 | -1.80 | (-2.46, -1.14) | <0.001 |
| max FAO OXPHOS | 17.85 | -0.19 | -1.68 | (-2.44, -0.91) | <0.001 | -0.18 | -1.53 | (-2.28, -0.78) | <0.001 | -0.19 | -1.64 | (-2.41, -0.88) | <0.001 |
| ***RPE Fatigability*** |  |  |  |  |  |  |  |  |  |  |  |  |  |
| ATPmax | 0.15 | -0.12 | -0.22 | (-0.35, -0.09) | 0.001 | -0.10 | -0.20 | (-0.33, -0.06) | 0.004 | -0.07 | -0.14 | (-0.27, -0.00) | 0.05 |
| max OXPHOS | 18.44 | -0.19 | -0.36 | (-0.50, -0.21) | <0.001 | -0.17 | -0.31 | (-0.46, -0.17) | <0.001 | -0.14 | -0.26 | (-0.41, -0.12) | <0.001 |
| max FAO OXPHOS | 17.85 | -0.20 | -0.38 | (-0.56, -0.21) | <0.001 | -0.18 | -0.34 | (-0.52, -0.17) | <0.001 | -0.18 | -0.34 | (-0.51, -0.17) | <0.001 |

Model 1 adjusted for age, sex, race, site/technician; Model 2 adjusted for Model 1 + body mass index; Model 3 adjusted for Model 2 + accelerometry-measured total activity count
**Abbreviations:** ATPmax (mM/sec); Max OXPHOS – Maximal Complex I&II Supported OXPHOS (pmol/(s\*mg)); Max FAO OXPHOS – Maximal Complex I&II Supported FAO OXPHOS (pmol/(s\*mg)); OXPHOS – oxidative phosphorylation; FAO – fatty acid oxidation; PFS – Pittsburgh Fatigability Scale; RPE – Borg Rating of Perceived Exertion; $\beta$ – standardized beta coefficient; $\beta$ - beta coefficient; CI – confidence interval

